# Leveraging past GMI vs A1c discordance markedly improves A1c prediction in a real-world cohort

**DOI:** 10.64898/2025.12.23.25342697

**Authors:** Giuliano Cruz, Ali Alqahtani, Nabeel Khan, Kate Hawke, Tom Elliott

**Affiliations:** Centre for Molecular Medicine and Therapeutics, British Columbia Children’s Hospital Research Institute, Vancouver, Canada; Department of Statistics, The University of British Columbia, Vancouver, Canada; BiomeHub, Florianopolis, Brazil; BCDiabetes, Vancouver, Canada; King Fahad Medical City, Riyadh, Saudi Arabia; Logan Endocrinology and Diabetes Service (LEADS), Logan Hospital, Meadowbrook, Australia; Faculty of Medicine, The University of British Columbia, Vancouver, Canada

**Keywords:** CGM, GMI, A1c

## Abstract

**Objective:** The glucose management indicator (GMI) is a widely used surrogate for A1c, but clinically significant GMI-A1c discordance is common. Here, we aim to develop a new GMI leveraging past discordance to improve accuracy in A1c prediction.

**Research Design and Methods:** This retrospective cohort study included 4,891 A1c records from 2,555 patients seen at BCDiabetes between 2021-Feb and 2025-Sep. Regression models adjusted for past discordance were trained on data from patients enrolled before 2025-Jan-01 and validated on those enrolled afterwards. Clinically significant discordance was defined as an absolute difference between GMI and A1c of ≥0.5%.

**Results:** In the validation cohort with 90-day CGM data, 39/256 (15.2%) patients with past discordance available showed clinically significant discordance between BCDiabetes GMI and A1c, compared to 101 (39.4%) patients with discordant Bergenstal GMI (relative risk [RR] 0.39, 95% CI 0.3—0.5, *P* < 0.0001). For BCDiabetes GMI, mean absolute discordance and Pearson correlation were 0.29% and 0.93, respectively, compared to 0.45% and 0.8 for the Bergenstal GMI. When including all validation patients regardless of availability of past discordance, BCDiabetes GMI showed discordance of 27.9% compared to 37% from Bergenstal GMI (RR 0.76, 0.69—0.83). Using 14-and-28-day CGM data, BCDiabetes GMI again showed reduced discordance compared to Bergenstal GMI with past discordance available (28-day RR: 0.54, 0.44—0.67; 14-day: 0.67, 0.56—0.80).

**Conclusions:** BCDiabetes GMI substantially reduces clinically significant discordance, especially when past discordance is available.

**Graphical abstract:** 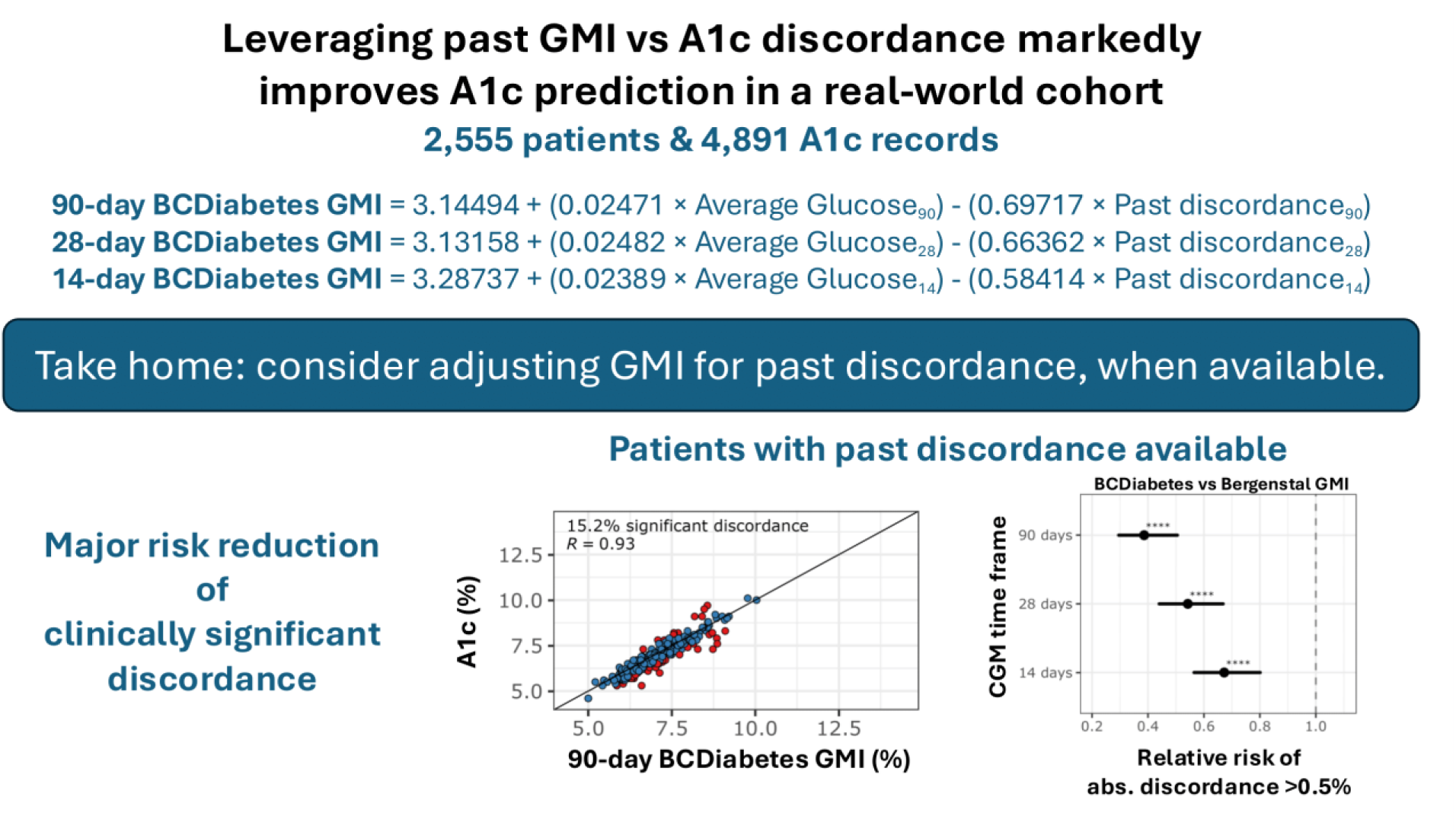

**Article Highlights:**

- **Why did we undertake this study?** GMI is a widely used surrogate for A1c, but clinically significant GMI-A1c discordance is common.
- **What is the specific question we wanted to answer?** Does accounting for past GMI-A1c discordance improve A1c prediction?
- **What did we find?** New GMI equations accounting for past GMI-A1c discordance markedly improved A1c prediction.
- **What are the implications of our findings?** Discordance-adjusted GMI may improve CGM-based decision-making.

**Twitter summary:** This work develops new GMI equations adjusted for past GMI vs A1c discordance, reducing the risk of clinically significant discordance (>0.5%) by over 60% when compared to the traditional GMI.”

## Introduction

The glucose management indicator (GMI) metric was formally introduced by Bergenstal et al in 2018 (1) as a term to replace “eA1C” and was derived from a linear regression equation based on mean CGM glucose in 548 individuals. Since then, many studies have evaluated GMI’s accuracy and limitations: GMI correlates with A1c but shows significant individual-level discordance. A 2024 review highlights that the relationship is nonlinear, and that GMI often underestimates A1c at higher values and overestimates A1c at low values (2). Reasons cited for this discordance include longer or shorter RBC life span, higher RBC glycation rates than average, and yet undefined biologic or genetic factors. In general, the negative correlation between A1c and GMI discordance (GMI minus A1c) is a consequence of variation in A1c that remains unexplained by average blood glucose with a population-level equation.

Recent work by Ajjan and colleagues proposes refined approaches (e.g., “uGMI” or red-cell dwell-time–adjusted metrics) aimed at personalizing A1c to account for inter-individual differences in erythrocyte glycation (3). Compared to A1c, the personalized A1c was more strongly correlated with Bergenstal GMI. Previous works also incorporated red cell lifespan into A1c correction (4). While valuable, these approaches do not focus on improving GMI, but change the definition of A1c itself. Another strategy incorporates nonlinearities into an updated GMI equation (uGMI) (5). The nonlinearity can be particularly relevant for individuals with extremely low or extremely high A1c. Yet, because uGMI remains a population-level function of average blood glucose, it cannot account for heterogeneity in the individual-specific relationship between A1c and average glucose.

Explicitly accounting for individual-level heterogeneity may require non-CGM measurements related to, e.g., red cell lifespan and glycation rates. However, for individuals who have measured A1c at least once under CGM use, departures from the population-level relationship between average blood glucose and A1c are reflected in the observed discordance between GMI and A1c. For an individual in which GMI consistently overestimates A1c, for example, adjusting future GMI predictions by subtracting the discordance (GMI-A1c) can therefore lead to improved predictions on average. Here, we aim to develop new GMI linear regression formulae, “BCDiabetes GMI” equations, which leverage individual past discordance to improve accuracy in A1c prediction.

## Research design and methods

### Study population

This retrospective cohort study included all patients with either Type 1 or Type 2 diabetes seen at BCDiabetes, a publicly funded pediatric and adult diabetes clinic in Vancouver, British Columbia between February 2021 and September 2025 – during the study period 10,751 individual patients were seen on at least one occasion. A database query was conducted to identify clients with at least one laboratory-measured A1c accompanied by CGM data immediately prior to the A1c measurement (A1c-CGM pairs). We included A1c-CGM pairs with CGM measurements for at least 70% of each time frame, considering 14, 28, and 90 days of CGM data paired with A1c (i.e., inclusion required CGM data available for at least 10 out of 14 days, 20/28, or 63/90).

This study was approved by University of British Columbia CREB # H25-01404-A001.

### Statistical analysis

All statistical analyses were conducted using R version 4.4.3. Baseline characteristics were summarized as medians (1^st^ and 3^rd^ quartiles) for continuous variables and frequencies for categorical variables. BCDiabetes GMI equations were developed using data from patients whose earliest A1c measurement date was before Jan 1, 2025 (development cohort). Validation included patients whose earliest measurement was on or after Jan 1, 2025 (validation cohort). We derived three equations, each using 90, 28, or 14 days of CGM measurements. For each equation, we employed linear regression using average glucose (mg/dL) and past discordance as predictors and A1c as the outcome. Past discordance was defined as Bergenstal GMI - A1c from a previous time point (i.e., a previous A1c-CGM pair). 90-day past discordance employs 90-day average glucose in the Bergenstal GMI equation (and analogously for 28- and 14-day past discordance). When unavailable, the past discordance was imputed using current average glucose. This is equivalent to using current average glucose as the only predictor when no past discordance is available. We quantified performance in the validation cohort using Pearson’s correlation coefficient, mean absolute discordance, and risk of clinically significant discordance. Clinically significant discordance was defined as an absolute difference between GMI and A1c of 0.5% or more. We compared the risk of clinically significant discordance between BCDiabetes GMI and Bergenstal GMI using Poisson regression with cluster-robust standard errors. To avoid pseudo-replication, all performance estimates are based on the single most recent A1cC-GM pair from each patient in the validation cohort. For greater precision, the final BCDiabetes GMI equations were re-estimated using all available data.

### Data and Resource Availability

The data sets analyzed in this study are not publicly available, due to privacy and ethical restrictions but may be available from the corresponding author upon reasonable request.

## Results

### Study population

This study included 4,891 pairs of A1c and CGM data from 2,555 individuals with Type 1 or Type 2 diabetes. Baseline characteristics are summarized in Table 1. The overall median A1c was 7 (6.4, 7.7) with a median age of 53 (33, 66) years. 300 (12%) individuals were under 18 years old and 1,200 (47%) were female. The median diabetes duration was 17 (8, 27) years.

**Table 1:** Baseline characteristics of the full cohort.

| Characteristic | N = 2,555 patients (4,891 A1c records) * |
| --- | --- |
| A1c | 7 (6.4, 7.7) |
| CGM type |  |
| Dexcom G6 or G7 | 1,702 (67%) |
| Libre 2 or 3+ | 853 (33%) |
| Age (years) | 53 (33, 66) |
| Age < 18 years | 300 (12%) |
| Female | 1,200 (47%) |
| Diabetes type |  |
| Type 1 | 1,480 (58%) |
| Type 2 | 1,075 (42%) |
| Diagnosis duration (years) | 17 (8, 27) |
| BMI (kg/m<sup>2</sup>) | 26 (23, 30) |
| Systolic BP (mmHg) | 125 (118, 130) |
| eGFR (mL/min/1.73m<sup>2</sup>) | 87 (64, 101) |
| Automated insulin delivery | 1,139 (45%) |
| Medication use |  |
| Basal insulin | 1,308 (51%) |
| Rapid insulin | 2,128 (83%) |
| GLP-1 receptor agonist | 778 (30%) |
| SGLT-2 inhibitor | 752 (29%) |
| Sulfonylureas | 25 (1.0%) |
| Metformin | 713 (28%) |

\*n (%); Median (Q1, Q3). Data shown combines development and validation cohorts.

### BCDiabetes GMI

To derive the BCDiabetes equations, we employed 3498 A1c-CGM pairs from 1721 patients in the development cohort. The resulting 90-, 28-, and 14-day BCDiabetes GMI equations are given below using units for glucose in mg/dL.

BCDiabetes GMI_90_ = 3.14494 + (0.02471 × Average Glucose_90_) - (0.69717 × Past discordance_90_)

BCDiabetes GMI_28_ = 3.13158 + (0.02482 × Average Glucose_28_) - (0.66362 × Past discordance_28_)

BCDiabetes GMI_14_ = 3.28737 + (0.02389 × Average Glucose_14_) - (0.58414 × Past discordance_14_)

A large past discordance means that Bergenstal GMI over- or underestimated A1c at a previous time and therefore BCDiabetes GMI is adjusted accordingly. If a patient has no past A1c measurement while under CGM use (e.g., just started using CGM, or not enough days of CGM immediately pre A1c), past discordance will not be available, so the following equations can be used instead.

GMI_90_ = 2.65471 + (0.02765 × Average Glucose_90_)

GMI_28_ = 2.81629 + (0.02675 × Average Glucose_28_)

GMI_14_ = 3.03613 + (0.02546 × Average Glucose_14_)

### Correlation between GMI and A1c

To validate the BCDiabetes equations, we employed the most recent pairs of A1c and CGM data from 834 patients in the validation cohort. Past A1c-CGM pairs, which are used to compute past discordance, were available for 362 patients. Fig. 1 shows the association between 90-day GMI and A1c. Overall, the 90-day BCDiabetes and Bergenstal GMIs showed Pearson correlation coefficients of 0.85 and 0.8, respectively. Among patients with past discordance available, the 90-day BCDiabetes GMI showed a Pearson correlation of 0.93, while Bergenstal GMI once again showed a correlation of 0.8. Similar results were observed for 28- and 14-day GMI (Supp. Fig. S1 and Supp. Fig. S2, respectively).

**Figure 1.**
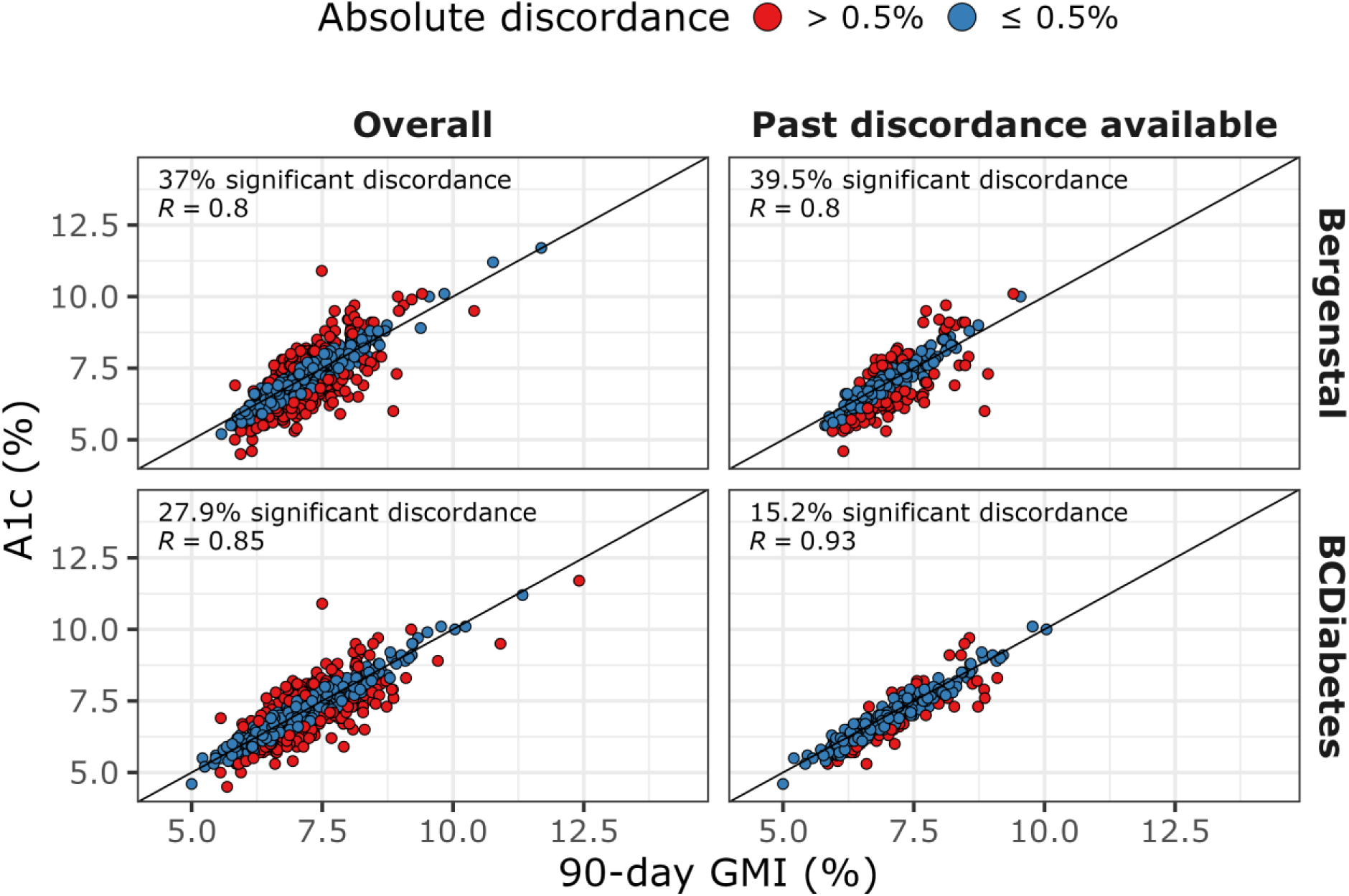
Scatter plots of 90-day GMI (%) and A1c (%) for Bergenstal (top) and BCDiabetes (bottom) GMIs. Overall (left) includes all patients in the validation cohort (n=834). Past discordance available (right) includes patients in the validation cohort with at least one past A1c-GMI pair available to compute past discordance (n=362). In all cases, each point represents the single most recent A1c-GMI pair from each patient. *R*: Pearson’s correlation coefficient.

### Risk of clinically significant discordance

Fig. 2 shows the mean absolute discordance and the percentage of patients with a clinically significant discordance (≥0.5%). BCDiabetes GMI led to lower discordance levels compared to Bergenstal GMI overall and, especially, among patients with past discordance available (Fig. 2A). For the 90-day GMI, the overall percentage of patients with clinically significant discordance dropped from 37.7% for Bergenstal to 27.9% for BCDiabetes (Fig. 2B). Bergenstal GMI was discordant for 39.5% of patients with past discordance available, compared to 15.2% for BCDiabetes GMI. As a result, the overall relative risk (RR) of clinically significant discordance was 0.76 (95% CI, 0.69 to 0.83, *P* < 0.0001), 0.8 (0.73 to 0.87, *P* < 0.0001), and 0.86 (95% CI, 0.8 to 0.92, *P* < 0.0001) for 90-, 28-, and 14-day CGM time frames, respectively, favoring BCDiabetes GMI (Fig. 3). Similarly, among patients with past discordance available, the RR was 0.39 (0.3—0.51, *P* < 0.0001), 0.54 (0.44—0.67, *P* < 0.0001), and 0.67 (0.56—0.8, *P* < 0.0001) for 90-, 28-, and 14-day CGM time frames, respectively.

**Figure 2.**
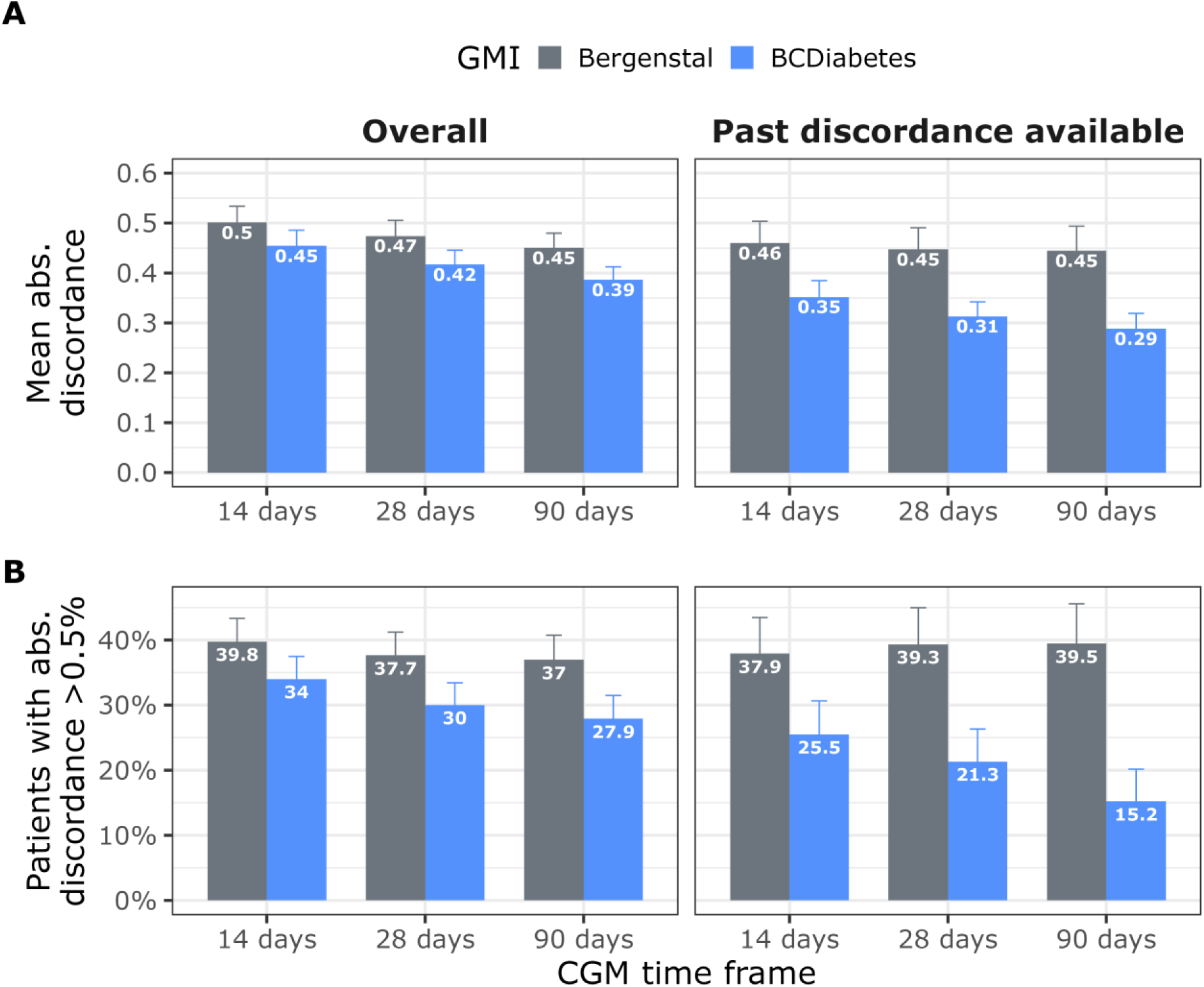
Absolute discordance between GMI and A1c in the validation cohort by CGM time frame. Clinically significant discordance rate is the proportion of patients with an absolute GMI vs A1c discordance greater than 0.5%. *Abs.*: absolute.

**Figure 3.**
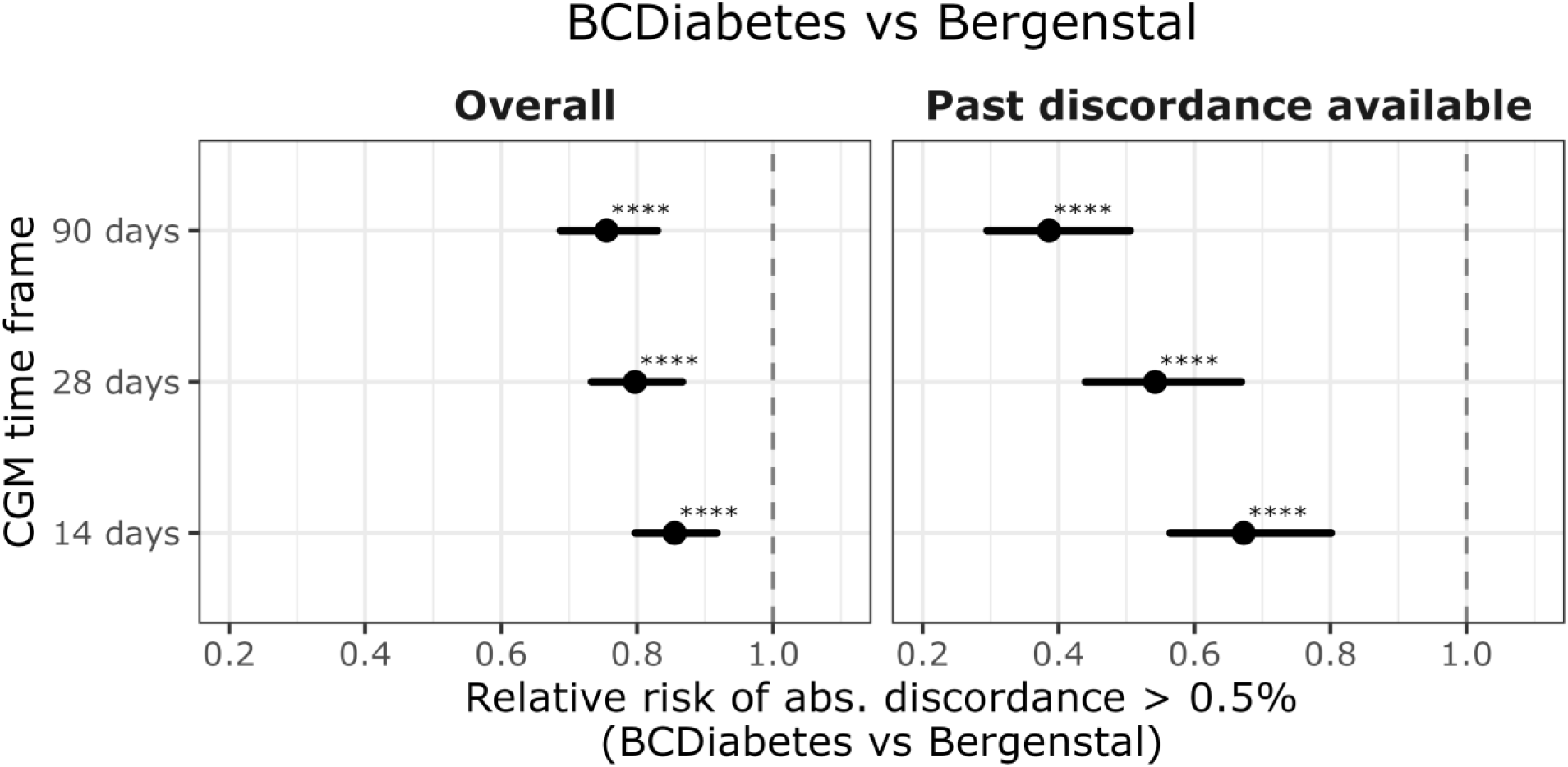
Relative risk of clinically significant discordance (>0.5%) between GMI and A1c in the validation cohort by CGM time frame. Values lower than 1 favor BCDiabetes GMI compared to Bergenstal GMI. *Abs.*: absolute.

There was no association between absolute discordance and the date gap between BCDiabetes GMI calculation and past discordance measurement, considering past discordance values measured up to 232 days before BCDiabetes GMI calculation (*P* ≥ 0.46 for all time frames, Supp. Fig. 3). When available past discordance values were ignored (i.e., using equation 2 for all patients), the mean absolute discordance for BCDiabetes GMI was slightly but consistently lower than for Bergenstal GMI (Supp. Fig. S4); in this case, BCDiabetes still led to a reduction in risk of clinically significant discordance, with a RR of 0.94 (0.89—0.9995, *P*=0.048), 0.95 (0.9—0.9985, *P*=0.043), and 0.98 (0.94—1.02, *P*=0.263) for 90-, 28-, and 14-day CGM time frames, respectively (Supp. Fig. S5).

## Discussion

This study uses real-world CGM and A1c data from a large pediatric and adult diabetes population to derive new GMI regression equations. The key difference from the original GMI equation is the adjustment for past discordance between Bergenstal GMI and A1c. When available, the past discordance captures the variation in patient-specific relationship between average blood glucose and A1c. This renders BCDiabetes GMI more personalized, without the need for additional, non-CGM measurements.

Overall, BCDiabetes GMI provides a moderate improvement over the Bergenstal GMI, achieving a Pearson correlation coefficient of 0.85 compared to the Bergenstal GMI’s 0.8; the overall percentage of patients with clinically significant discordance (absolute difference between GMI and A1c of 0.5% or more) dropped from 37.0% with Bergenstal GMI to 27.9% with BCDiabetes, a relative risk reduction of 25%.

When past discordance is available, BCDiabetes GMI strikingly reduces discordance compared to the Bergenstal GMI. Among patients with known past discordance measured up to 232 days before BCDiabetes GMI calculation, the Pearson correlation coefficient increased to 0.93, reducing the percentage of patients with clinically significant discordance from 39.5% to 15.2%, a relative risk reduction of 62%. As a result, for patients with past discordance information, BCDiabetes GMI can prevent 6 out of 10 clinically significant discordance events from Bergenstal GMI.

Without any past discordance information, BCDiabetes GMI showed a relative risk reduction of about 5% for the 90- and 28-day CGM time frames. This improvement is likely in part due to the current study’s real-world design and larger sample size: Bergenstal GMI was derived from 548 individuals in 4 separate clinical trials with an average CGM duration of 48 days (1). The BCDiabetes GMI was derived with 4891 A1c-CGM pairs from 2555 real-world patients with an average CGM duration of 13, 26, and 81 days for the 14-, 28-, and 90-day equations, respectively.

These new GMI equations are unlikely to change clinical decision-making for patients with consistently optimal or inadequate A1cs. For patients with ideal or optimal A1c for whom GMI has been discordantly high, the adjusted BCDiabetes GMI will provide reassurance to both patients and clinicians. For patients with sub-optimal A1c for whom the simple BCDiabetes GMI is relatively flattering, use of the adjusted BCDiabetes GMI may support intensification of treatment without concurrent A1c measurement.

A potential weakness in this study is its single site design - this is mitigated by the observation that 70% of patients live outside the city of Vancouver and 35% live in Canadian provinces other than British Columbia. While the A1c method was not tracked, all A1c measurements were performed at accredited commercial laboratories.

The simple and discordance-adjusted BCDiabetes GMI equations represent promising advances in glucose monitoring, potentially leading to increased acceptance of GMI by both clinicians and patients, improved clinical decision-making, and decreased reliance on laboratory A1c.

## Data Availability

The data sets analyzed in this study are not publicly available, due to privacy and ethical
restrictions but may be available from the corresponding author upon reasonable request.

## Acknowledgments

The authors acknowledge all BCDiabetes patients and staff. The authors thank Dr. Keegan Korthauer for helpful comments and discussion of the work.

## Author contributions

G.C., A.A., and T.E. performed formal analysis and wrote the first draft of the manuscript. G.C., A.A., N.K., K.H., and T.E contributed to the discussion and reviewed and edited the manuscript. N.K., A.A, and T.E. acquired the data. All authors approved the final version of the manuscript. T.E. is the guarantor of this work and, as such, had full access to all the data in the study and takes responsibility for the integrity of the data and the accuracy of the data analysis.

## Supplement

## Supplementary Figures

**Supp. Fig. S1.**
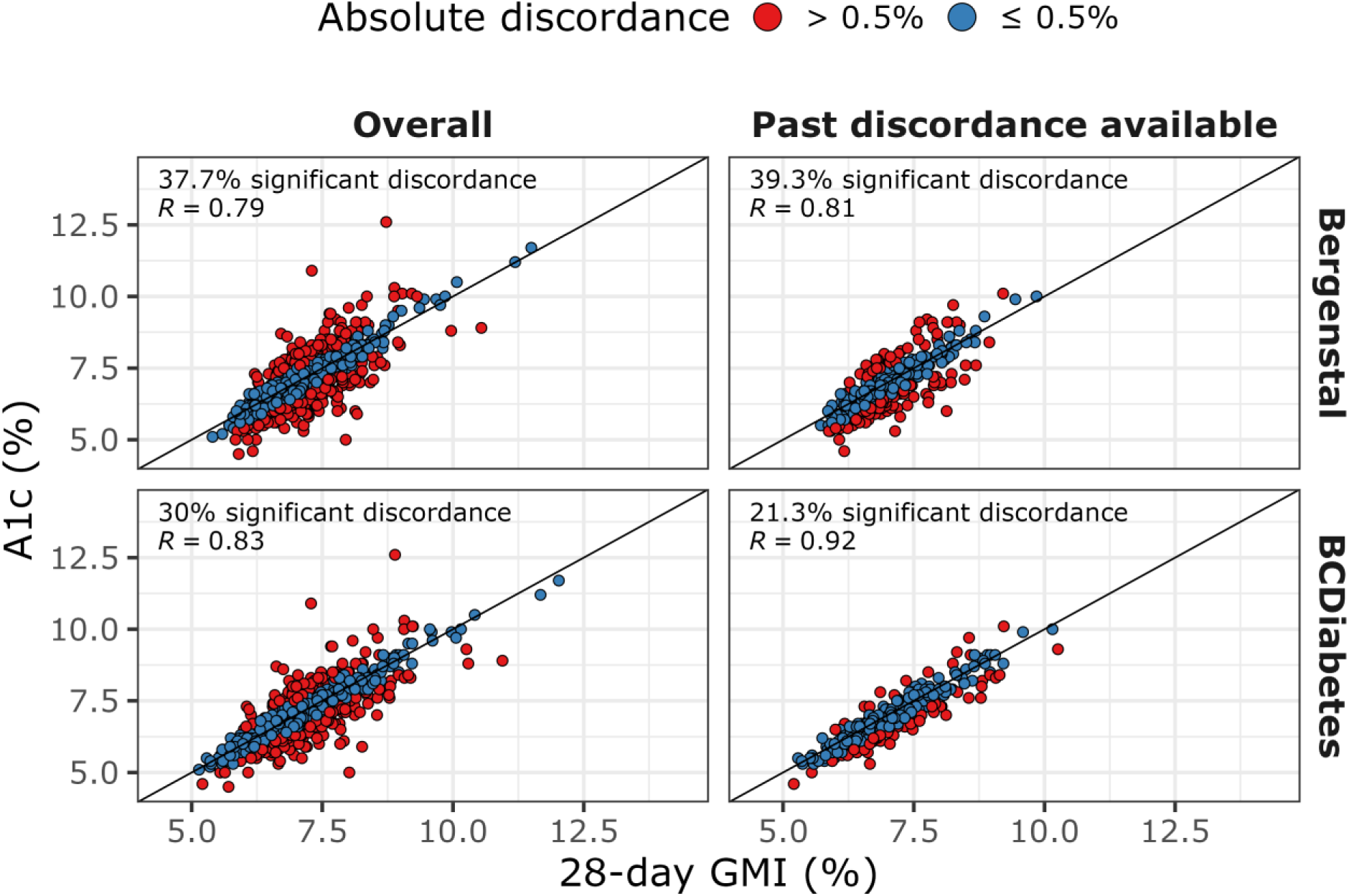
Scatter plots of 28-day GMI (%) and A1c (%) for Bergenstal (top) and BCDiabetes (bottom) GMIs. Overall (left) includes all patients in the validation cohort (n=834). Past discordance available (right) includes patients in the validation cohort with at least one past A1c-GMI pair available to compute past discordance (n=362). In all cases, each point represents the single most recent A1c-GMI pair from each patient. R: Pearson’s correlation coefficient.

**Supp. Fig. S2.**
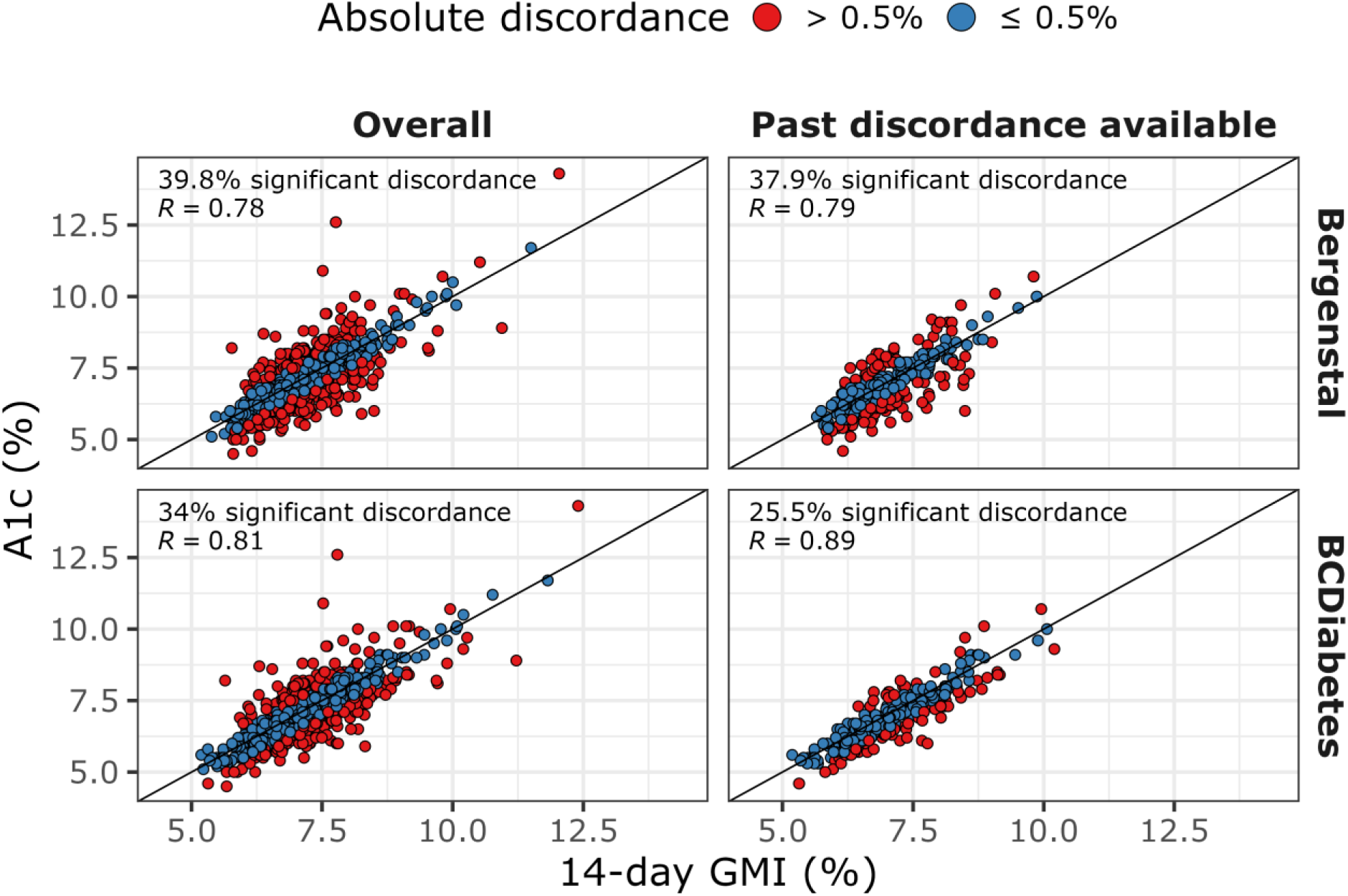
Scatter plots of 14-day GMI (%) and A1c (%) for Bergenstal (top) and BCDiabetes (bottom) GMIs. Overall (left) includes all patients in the validation cohort (n=834). Past discordance available (right) includes patients in the validation cohort with at least one past A1c-GMI pair available to compute past discordance (n=362). In all cases, each point represents the single most recent A1c-GMI pair from each patient. R: Pearson’s correlation coefficient.

**Supp. Fig. S3.**
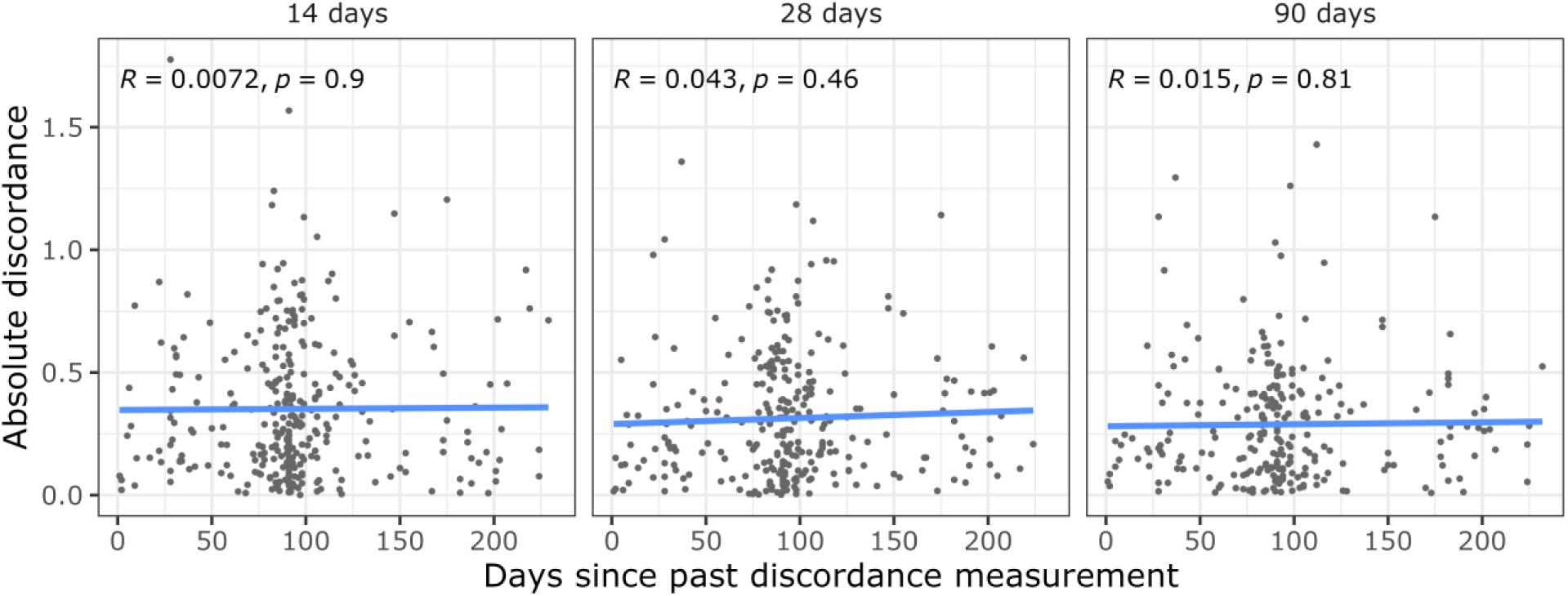
Absolute discordance between BCDiabetes GMI and A1c by time (days) since past discordance measurement in the validation cohort among patients with at least one past A1c-GMI pair available.

**Supp. Fig. S4.**
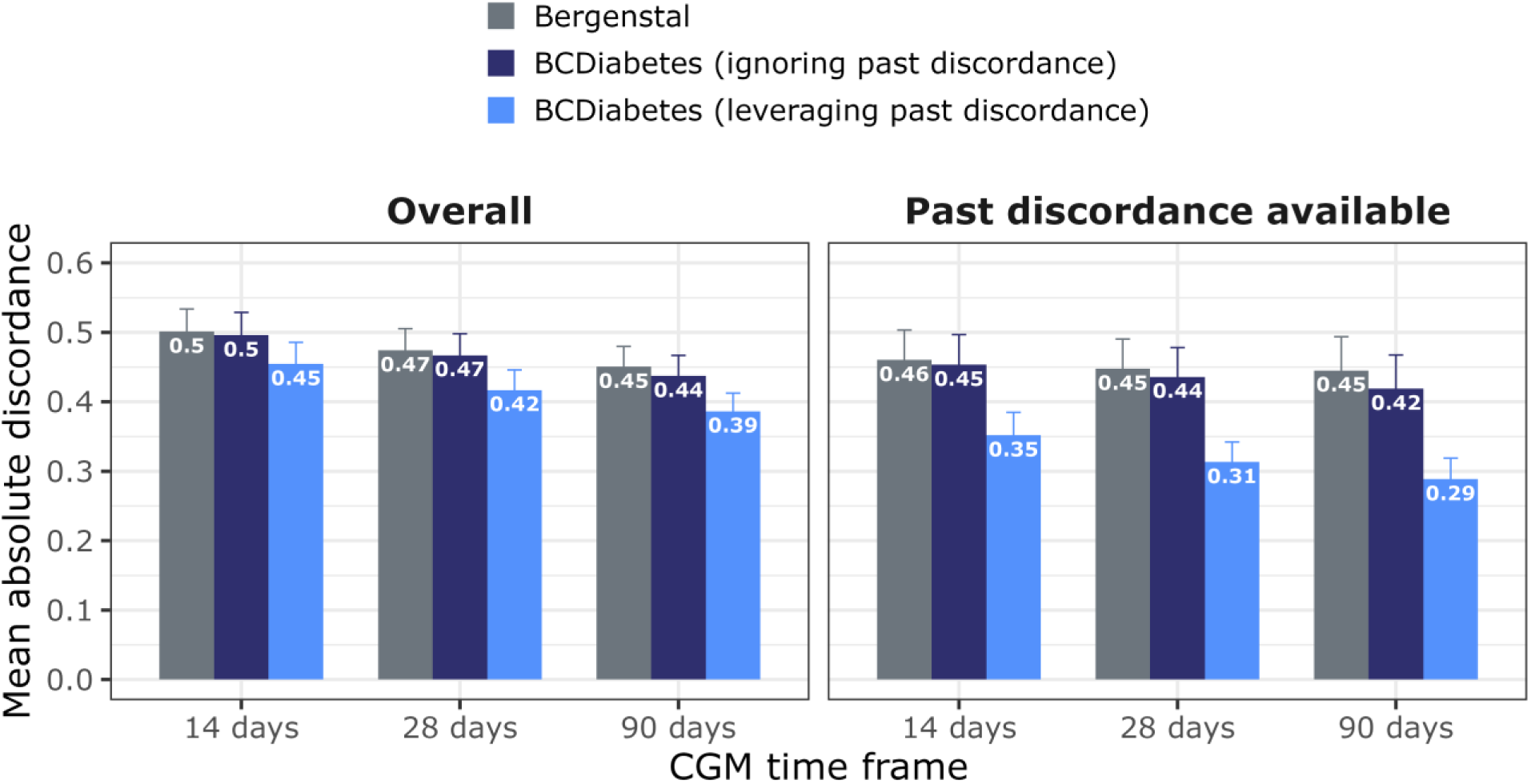
Absolute discordance between GMI and A1c in the validation cohort by CGM time frame, including BCDiabetes GMI, Bergenstal GMI (leveraging past discordance when available), and BCDiabetes GMI ignoring any past discordance values.

**Supp. Fig. S5.**
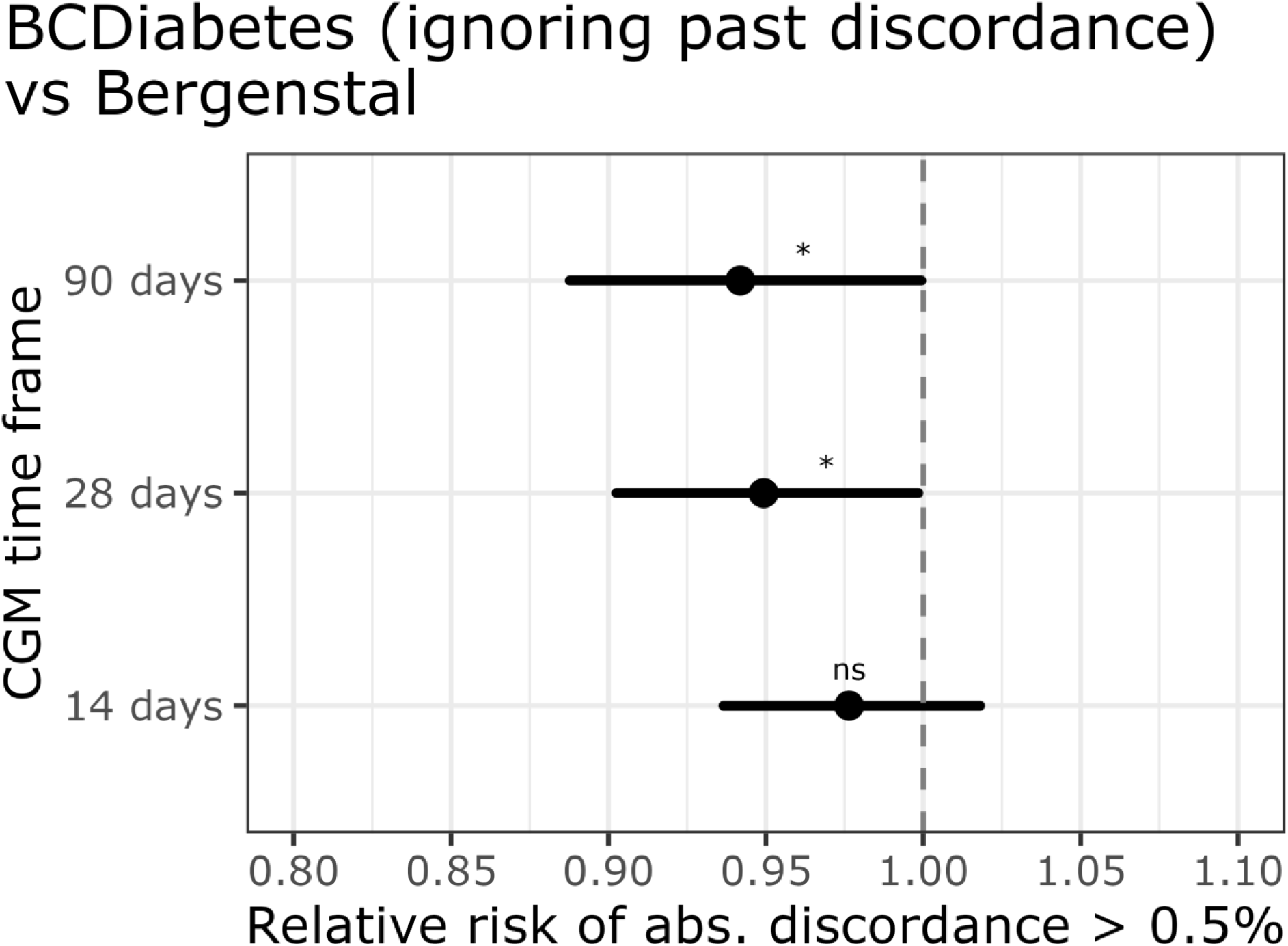
Relative risk of clinically significant discordance (>0.5%) between GMI and A1c in the validation cohort by CGM time frame. Values lower than 1 favor BCDiabetes GMI (ignoring past discordance) compared to Bergenstal GMI. Abs.: absolute.

